# A Pilot Study of Generative AI video for Patient Communication in Radiology and Nuclear Medicine

**DOI:** 10.1101/2024.11.06.24316678

**Authors:** MK Badawy, K Khamwan, D Carrion

## Abstract

**Purpose:** Effective patient communication for radiation imaging procedures is critical, especially for patients with literacy issues or language barriers. Generative Artificial Intelligence (GenAI) provides a new approach for creating personalised, multilingual patient education materials. This pilot study evaluates the effectiveness of GenAI, specifically using HeyGen, in creating personalised patient information videos in the Thai language.

**Methods:** We created an avatar of a medical physicist using HeyGen. Two English health information scripts on nuclear medicine and radiology were translated into Thai using HeyGen’s translation tool, and videos were created with the avatar delivering the content in Thai. Thirteen native Thai-speaking medical physicists and postgraduate students evaluated the videos using a 5-point Likert scale, focusing on translation accuracy, naturalness of delivery, and usefulness as a patient education tool. The Bilingual Evaluation Understudy scoring system was used to assess translation quality objectively.

**Results:** Both videos received high median scores for translation accuracy (median of 4.0). The BLEU scores were 0.56 and 0.66, indicating good translation quality. Participants reported minor issues with formal language and unnatural phrasing, but overall found the videos understandable and useful. Feedback suggested improving the naturalness of the avatar’s delivery to increase relatability.

**Conclusions:** Our pilot study shows that GenAI can effectively create and translate personalised patient information videos into Thai, helping to bridge communication gaps in radiation-related procedures. While minor issues remain, the findings indicate that tools like HeyGen could significantly improve patient communication, particularly for those who face language barriers.

## 1 Introduction

Effective communication is essential in healthcare, as it helps individuals understand and manage their health conditions, leading to better engagement and participation in their care [1,2]. In the realm of imaging and radiation treatment, where the risk of radiation can cause anxiety, providing clear and accessible patient information is particularly important [3]. Using videos to deliver patient information has been shown to enhance patients’ understanding of their conditions and treatment plans, improving communication between patients and healthcare providers [4–6]. Whilst videos are generally well-received and can reduce patient anxiety [7], many patients prefer direct oral communication for its clarity and personal touch, making it more relatable and engaging than standard videos [8].

However, barriers like illiteracy and language differences can hinder effective communication in imaging and radiation treatment. Patient illiteracy hampers understanding, treatment compliance, and access to care [9,10]. Moreover, even literate patients may face challenges when they and their healthcare providers do not share a common language, which is not uncommon in diverse healthcare settings. While interpreters can help overcome language barriers [11], access to them is not always feasible. Automated translation applications have been suggested as alternatives, but concerns exist regarding their accuracy, particularly in translating complex medical terminology [12,13].

Generative artificial intelligence (GenAI) offers a novel solution in creating synthetic personalised communication that can simulate a conversational experience tailored to patients’ individual needs. GenAI has been used to make synthetic learning videos for general education [14], achieving results comparable to traditional video formats. Our study explores GenAI’s unique capability to simulate a personalised, conversational experience that can engage patients directly. HeyGen, for instance, can generate high quality avatars and translate content into numerous languages. It has been used in medical settings to create synthetic patients to assist doctors with communicating complex medical information around sensitive topics medical [15] and translate patient information in ophthalmology [16]. No study to date has explored its potential in providing personalised oral communication via video directly to patients in the context of radiation imaging procedures.

Our pilot study assesses whether GenAI can create personalised, avatar-based patient information videos in Thai, addressing both language barriers and the need for patient-centred communication. By utilising avatars that simulate oral communication, this approach aims to deliver a more personalised experience than standard translation applications, enhancing patient understanding and engagement. The findings from this pilot will inform future expansions, including adding other languages and the broader application of GenAI-generated, avatar-driven content in patient settings.

## 2 Methods

### 2.1 Ethical considerations

No patient or sensitive information was collected during the study, and no ethical approval was necessary since the video featured one of the authors. Informed consent was obtained from both the author appearing in the video and the survey participants. As required by HeyGen, a separate consent video was recorded and uploaded to authorise the avatar’s generation.

### 2.1 Creation of the avatar

Following HeyGen’s instructions as of 1st October 2024 (Appendix A), we recorded a 3-minute 4K video of the medical physicist standing two metres from the camera in front of a green screen. To ensure the avatar accurately replicated the author’s appearance and mannerisms, the author kept eye contact, spoke naturally, and kept their hands below chest level and out of frame to focus on facial expressions and avoid awkward movements.

We uploaded the main footage and consent video to the HeyGen AI platform. Using the version active on 1st October 2024, the platform processed the videos to capture the author’s facial expressions, lip movements, and voice characteristics. This resulted in an avatar closely replicating the author’s appearance and mannerisms to be used for video generation.

### 2.3 Creation of the AI Voice

The voice for the AI avatar was recorded separately, with the medical physicist instructed to speak slowly and with exaggerated pronunciation. The recording focused on capturing phrases relevant to the project’s content, ensuring that key vocal characteristics such as tone, pitch, and clarity were well defined. The voice recording was then uploaded to HeyGen’s AI Voice service, where it was used to generate an AI voice that accurately reflected the individual’s natural speaking patterns.

### 2.4 Development of Health Information Content

The author, a Medical Physicist and a Radiation Safety Officer with more than ten years of experience, developed two health information scripts in English:

1. Nuclear Medicine Script: Information about the safety precautions required following radionuclide therapy.
2. Radiology Script: Instructions for patients to check for radiation-induced skin injury after undergoing complex interventional radiology procedures.

These scripts were crafted to be easily understandable by patients and are available as supplementary material for reference. The scripts can be made available by request to the authors.

### 2.5 Translation and video generation

We used HeyGen’s “Create Video” function to create two personalised patient communication videos. The English script was entered into the “Script” section and translated into Thai using HeyGen’s built-in translation tool (original English and translated Thai scripts can be provided by request to the authors). We selected the previously generated AI voice and set the language option to “Thai - Default Locale” to ensure appropriate linguistic and regional alignment. The custom avatar created earlier was chosen as the presenter.

A CT scanning room background was added to enhance visual context, and we applied the “Remove Green Screen” option to integrate the avatar seamlessly into the background. The video was then generated (Figure 1).

**Figure 1.**
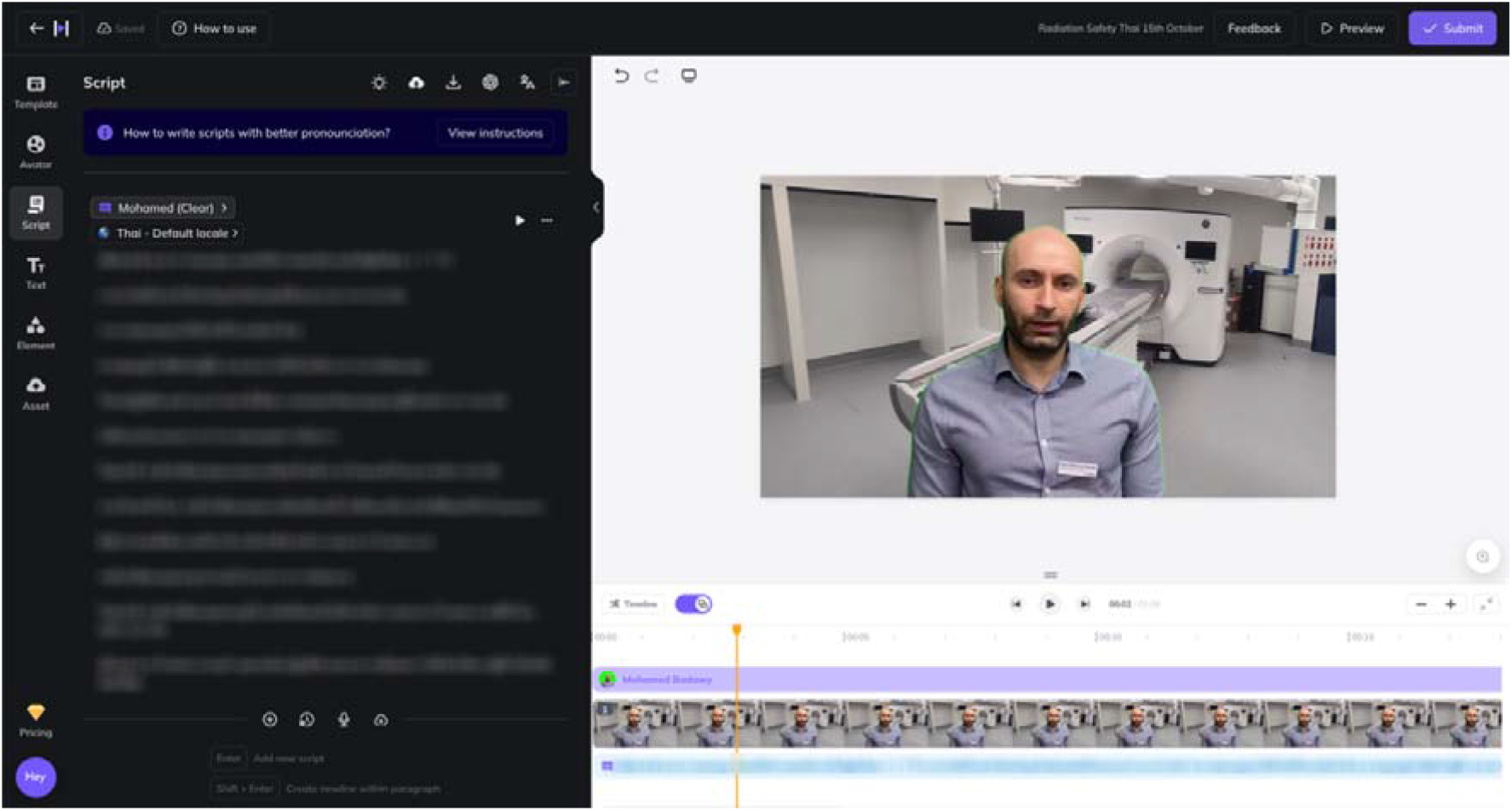
HeyGen video creation section featuring a translated Thai script (left), video preview window (right), and the timeline for syncing script with visuals (bottom). The “Submit” button (top right) allows for video generation.

After the video was generated, we downloaded the video in MOV format with subtitles enabled to improve accessibility (Figure 2). Due to minor green screen artifacts, suspected because of suboptimal lighting conditions, we used the 3D Keyer effect in DaVinci Resolve 18 to eliminate residual green screen remnants and exported the video in MP4 format. The final video was uploaded to YouTube as an unlisted link and incorporated into the survey to evaluate the GenAI-generated video.

**Figure 2.**
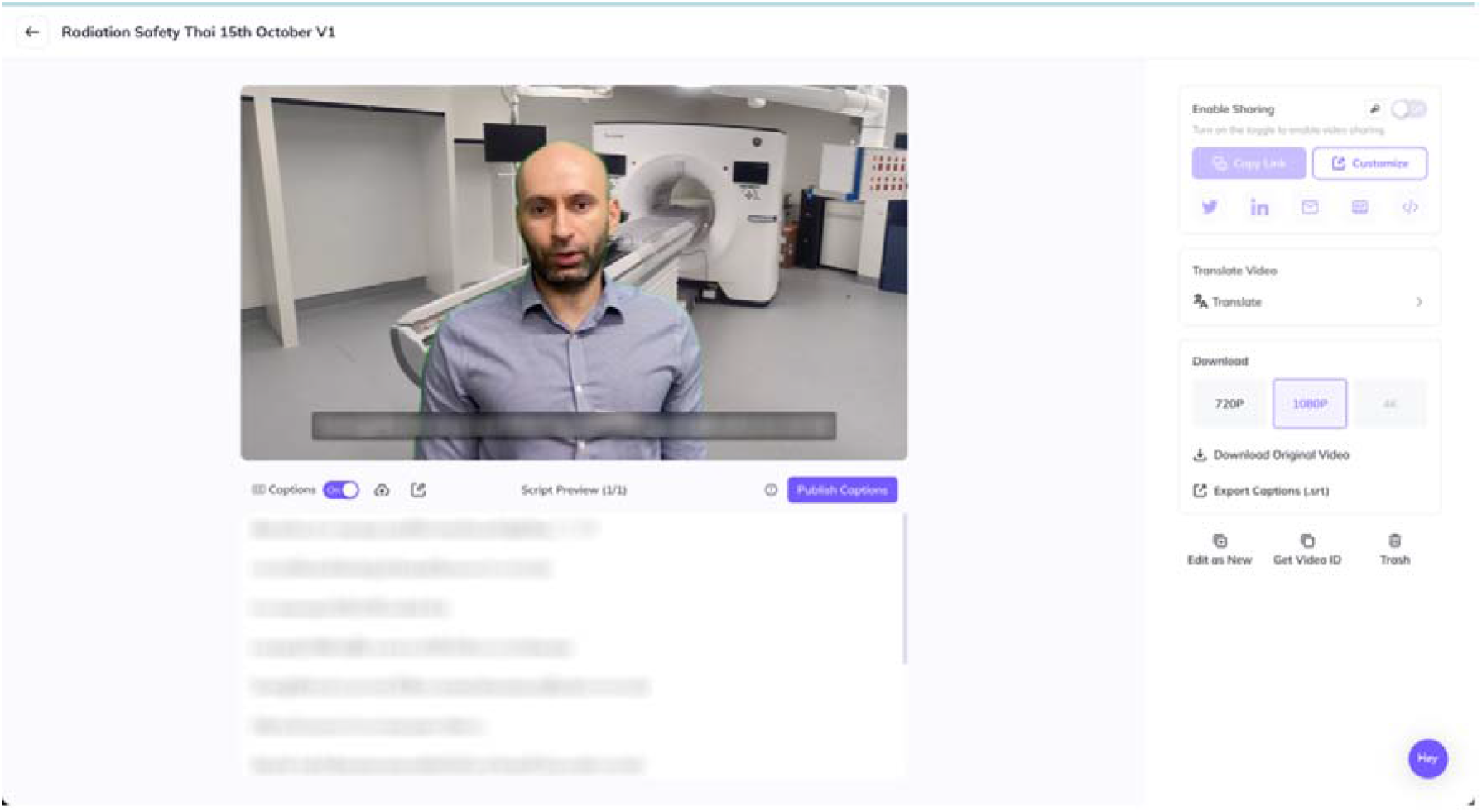
Interface to download video with the option of selecting captions for inclusion in the video.

### 2.6 Participant recruitment and evaluation

We recruited native Thai speakers rank the quality of the translations and the effectiveness of the videos. Participants were postgraduate students in medical physics and clinical medical physicists, to ensure familiarity with radiation safety risk communication and the nuances in radiology and nuclear medicine medical physics. All participants watched both videos together in the same room and they completed a survey via an online Google form based on specified criteria.

The criteria assessed on a Likert scale, 1-5, for each video, were:

1. Accuracy of translation
2. Quality of accent
3. Naturalness of the delivery
4. Naturalness of the avatar
5. Accuracy of medical terminology
6. Accuracy of scientific terminology
7. Usefulness in patient information space
8. Impact to patient access to radiation information
9. Likelihood of using this tool

Participants rated criteria 1–9 on a 5-point Likert scale (1 = Poor, 5 = Excellent). For criterion 10, participants provided open ended feedback highlighting specific issues.

### 2.7 Data Analysis

Quantitative data from the Likert scale ratings were analysed using descriptive statistics, including median and range for each criterion. We used the Wilcoxon Signed-Rank test to compare the paired participant ratings for each criterion across the two videos and the Mann-Whitney U test to compare the rating distributions across professional backgrounds.

Qualitative feedback from participants was thematically analysed to identify common issues, strengths, and suggestions for improvement. This involved coding responses, grouping them into themes, and interpreting the findings in the context of the study’s objectives.

### 2.8 Objective Evaluation of Translation Quality

To objectively assess the quality of the machine translations produced by the GenAI model, we used the Bilingual Evaluation Understudy (BLEU) scoring system. BLEU is a standardised metric that evaluates the similarity between machine-translated text and one or more high-quality human reference translations, providing a quantitative measure of translation accuracy [17]. The BLEU scores provided an objective metric for evaluating the translation quality, with scores closer to 1 indicating a higher degree of similarity to the human translations. A score greater than 0.5 is generally regarded as good.

A native Thai speaker specialising in medical physics and radiation safety was used to create accurate reference translations of the original English scripts to ensure the reference translations were high quality and contextually appropriate.

The BLEU scores were calculated by comparing the HeyGen tool translated texts to the corresponding human reference translations. We used Python 3.10 and nltk version 3.9.1 package to calculate the BLEU scores.

## 3 Results

The evaluation included native Thai speakers with medical imaging and radiation safety backgrounds, who rated two GenAI-generated videos on criteria such as translation accuracy, medical terminology, accent quality, and avatar delivery naturalness. Both videos scored a median of 4.0 on translation accuracy and quality of accent, with Video 1 showing a statistically significant advantage in medical terminology accuracy (p = 0.045). No significant differences were found in criteria of naturalness of delivery or usefulness in patient education. BLEU translation scores were 0.56 and 0.66 for Videos 1 and 2, respectively, suggesting good translation accuracy. Qualitative feedback highlighted minor issues with formal language and awkward phrasing in both videos, though participants found them generally understandable and helpful for patient communication.

### 3.1 Participant demographics

Most participants were of Thai backgrounds (13), with a smaller representation of Burmese (2)native speakers. Only the Thai participants were analysed. The group included individuals with various experience levels, from entry-level (0 years) to more seasoned professionals (up to 15 years). Participants’ ages ranged from 23 to 42, with a representation more skewed to the female gender.

**Table 1.** Summary of Participant Demographics by professional background.

| Professional Background | n | Gender (F%) | Age (years) (median, range) | Experience (years) (median, range) |
| --- | --- | --- | --- | --- |
| Medical Physicist | 7 | 71 | 27.0 (27-42) | 1.0 (0.0-15.0) |
| Postgraduate Student | 6 | 67 | 24.0 (23-31) | 1.5 (0.0-5.0) |

### 3.2 Quantitative evaluation of the videos

The results from the analysis are presented in Figure 3. Overall, both videos received similar median scores across most criteria. The lowest median scores were observed in the “Naturalness of delivery,” where Video 1 scored 3.5 (range 2.0-5.0) and Video 2 scored 3.0 (range 2.0-5.0). The highest median scores were in the “Usefulness in patient information,” with Video 1 receiving 5.0 (range 3.0-5.0) and Video 2 receiving 4.0 (range 3.0-5.0). A statistically significant difference was found in the accuracy of medical terminology, where Video 1 performed better than Video 2 (p-value = 0.046). This suggests that while both videos are generally effective, the terms used in Video 1 may have been translated more naturally into Thai. The sensitivity of the Wilcoxon Signed-Rank test with small sample sizes should be considered when drawing conclusions.

**Figure 3.**
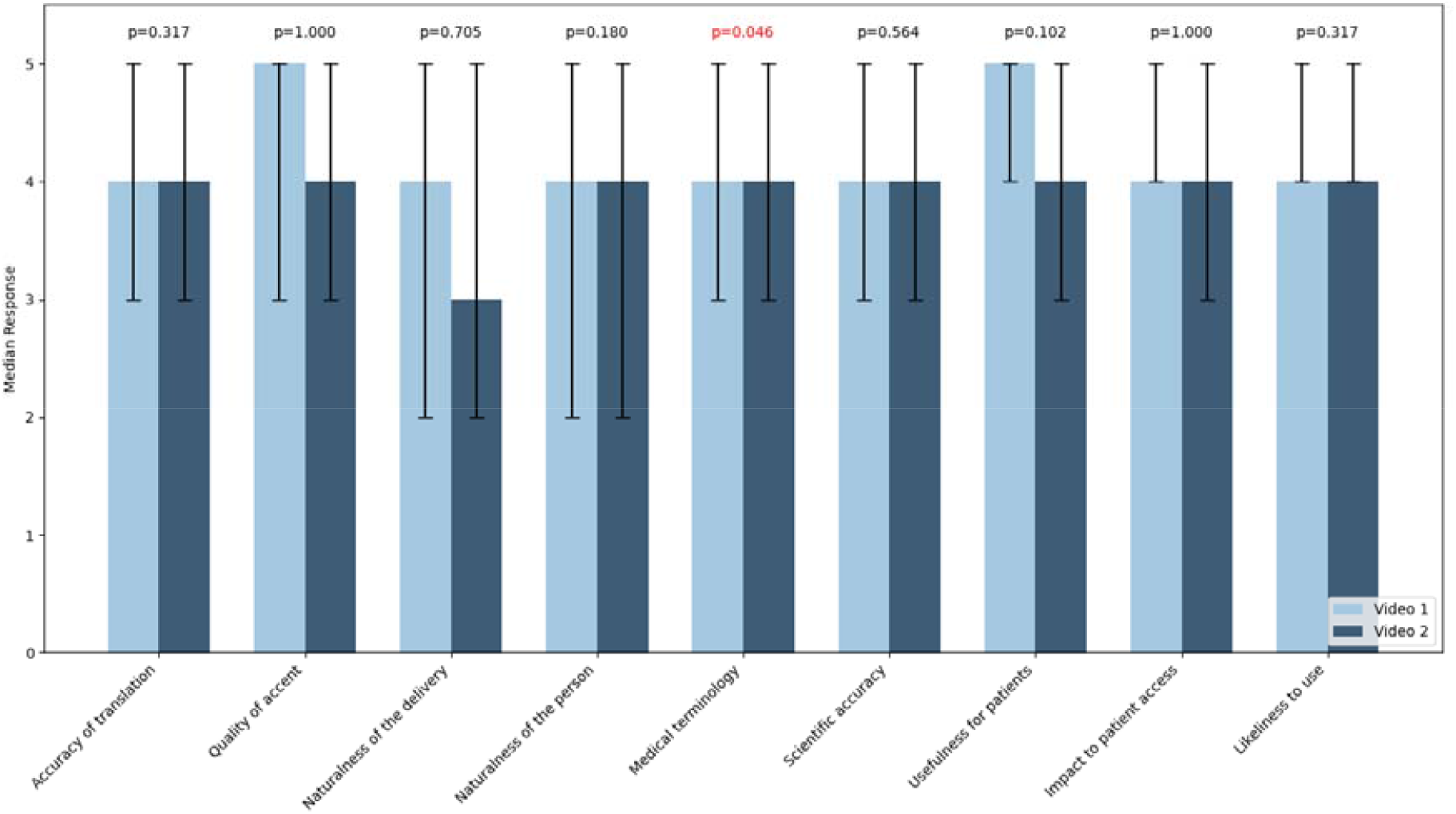
Comparison of two videos by evaluation criteria with significant value highlighted.

Both Medical Physicists and Students provided similar ratings across most criteria, with high median scores ranging from 3 to 5 (Figure 4). The largest discrepancy was in the “Naturalness of delivery,” where Students reported a slightly lower median score of 3.5 compared to Medical Physicists’ 4.0. No statistically significant differences between the two groups were revealed for any of the criteria (p□>□0.05), indicating consistent evaluations across all categories.

**Figure 4.**
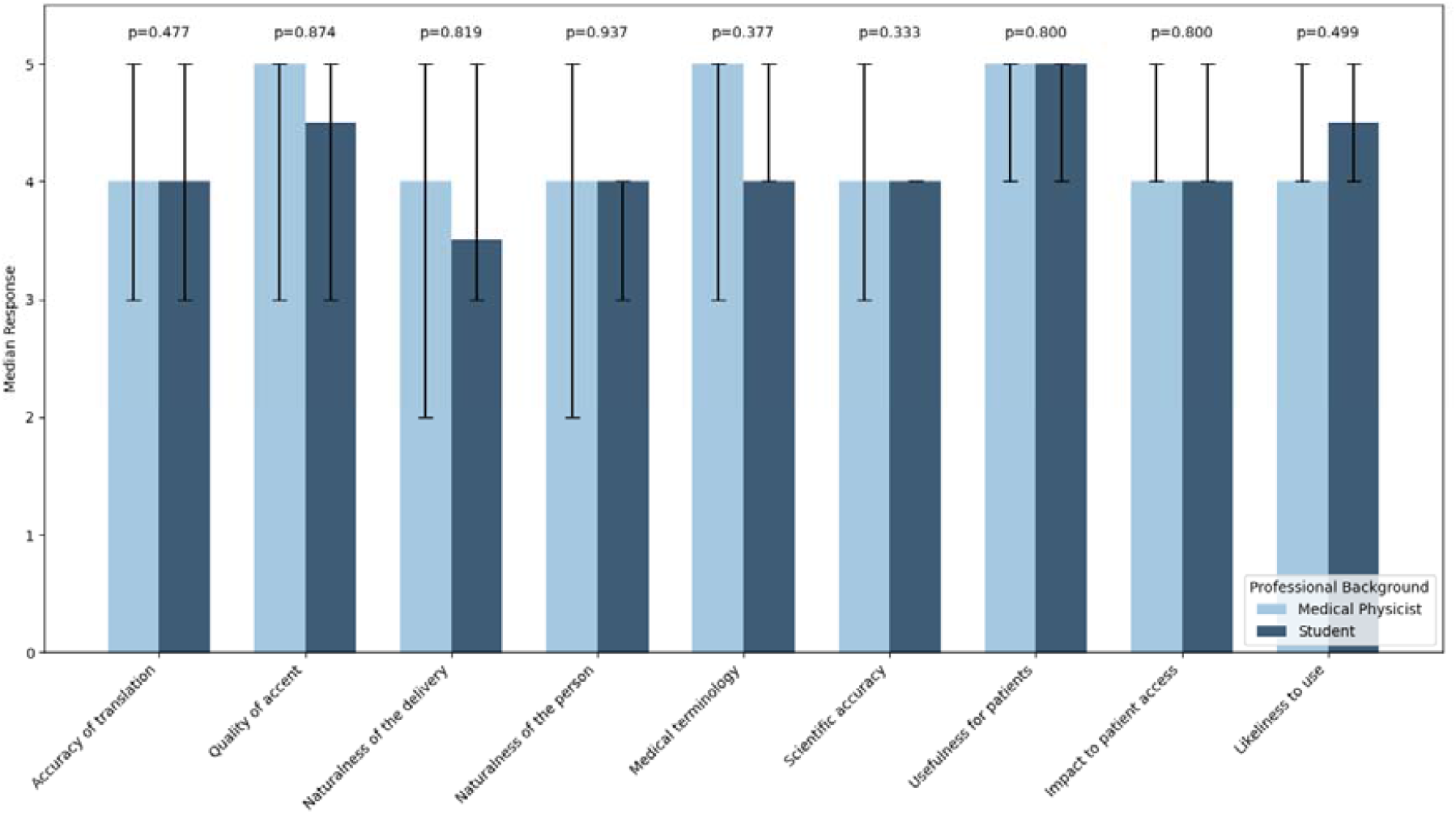
Comparison of ratings by professional background and criteria

### 3.3 Objective Evaluation of Translation Quality

As shown in Table 4, Video 1 received a BLEU score of 0.56. In contrast, Video 2 had a slightly higher score of 0.66, indicating that Video 2’s translation was marginally more accurate according to the BLEU metric. However, participants rated both videos equally, with a median rating of 4.0 for Accuracy of Translation. This suggests that the difference in BLEU scores did not significantly impact their perception of translation quality.

**Table 4.** Summary of BLEU scores and participant median rating.

| Video | BLEU Score | Participant Median Rating |
| --- | --- | --- |
| Video 1 | 0.56 | 4.0 |
| Video 2 | 0.66 | 4.0 |

Due to the small sample size of BLEU scores (n=2), performing a meaningful correlation analysis using standard statistical methods such as Pearson or Spearman correlation was not feasible. These methods require a larger set of data points to establish robust relationships between variables, in this case, only two BLEU scores (one for each video).

### 3.4 Qualitative Feedback from Participants

Participants identified key areas where translations in both videos were incorrect or unnatural (Table 5). In Video 1, the use of “therapy” was flagged, with “treat” suggested as a more appropriate term for “treatment.” Some words had more precise synonyms, and specific sentences sounded unnatural to native speakers. In Video 2, unnatural grammar and word order impacted the translation’s fluidity; for example, “maybe happen” would more naturally be expressed as “possible” in colloquial Thai. Despite these issues, both videos were generally understandable. Suggestions for improvement included using less formal language, enhancing the naturalness of the avatar’s movements (especially for older viewers) and consulting native speakers to refine grammar and phrasing for more conversational Thai.

**Table 5.** Summary of video and issues identified by participants.

| **Video 1** | **Video 2** |
| --- | --- |
| "therapy" < "treat"<br>A little bit too formal but understandable.<br>A little bit stiffness might be uncomfortable for geriatrics.<br>The word for urine, HeyGen AI translates this as "Pas-sa-wa," which is technically correct, it sounds unnatural. In our conversation, we typically use a more blended pronunciation, "Passawa," which sounds much more natural. | Grammar and the word order are not quite natural. Might need more advice from native speaker. |
| There are some words that have a more proper synonym than the one in the video, some sentences are wrong in order so it's kind of strange but still understandable, and some pause after sentence is incorrect rhythm. | "Maybe happen" translated correct but in native usage we use it as it means "possible" |

## 4 Discussion

### 4.1 Summary of Key Findings

Our study evaluated the effectiveness of GenAI, specifically HeyGen, in producing personalised, multilingual patient communication videos for imaging and treatment involving radiation by assessing translation accuracy, naturalness, and usefulness. The findings indicate that HeyGen can effectively generate such videos, receiving favourable subjective participant feedback and achieving good translation accuracy as measured by BLEU scores. While translations were largely accurate, some issues related to complexity and medical terminology were noted, suggesting that further refinement is needed for broader application. Participants found the videos helpful as educational tools but mentioned challenges related to the naturalness of the avatar’s delivery.

### 4.2 Interpretation of Findings

Both videos demonstrated good translation accuracy and were considered useful for patient communication in radiation imaging context. Participants rated both videos highly for translation accuracy, which aligned with BLEU scores, indicating reasonable agreement with their perceptions. However, issues were noted regarding the naturalness of specific translated phrases, the formal tone of the language, and awkward phrasing that affected the fluidity of the spoken language. These challenges highlight the complexities of translating medical information into languages with different grammatical structures and nuances, especially when dealing with specialised medical terminology.

Some participants observed that the avatar’s delivery appeared slightly stiff, potentially impacting effectiveness in real-world healthcare settings where relatability and engagement are crucial, particularly for older or less tech-savvy audiences. This stiffness may be attributed to the initial video recording methods. Adjusting the recording techniques could make the avatar appear more natural and friendly.

Technical challenges were also encountered. The initial audio recording for the AI avatar was unusable due to low volume, necessitating re-recording with more controlled delivery and deliberate speech patterns. This emphasises the importance of high-quality input to ensure accurate voice modelling by GenAI systems. Additionally, while HeyGen performed well overall, the green screen removal required post-production enhancement using external software (DaVinci Resolve 18). Enhanced green screen keying options within HeyGen could streamline this process, reducing reliance on additional editing tools.

### 4.3 Comparison with Existing Literature

Our findings are consistent with those reported in a recent exploratory study [16] that also assessed the use of HeyGen for generating patient information videos across multiple languages. In that study, a Danish patient information video on eye drop usage was translated into six languages: English, German, French, Polish, Arabic, and Turkish. These vary in linguistic distance from Danish. The translations were evaluated by ophthalmologists proficient in Danish and the target languages. The study found that while translations into closely related languages were highly accurate, translations into remotely related or unrelated languages contained several inaccuracies and unnatural phrasing. Despite these issues, the overall messages remained understandable due to the visual context provided by the videos.

Our results and those of the previous study highlight the potential of GenAI tools like HeyGen to produce comprehensible patient information across languages, albeit with translation accuracy and naturalness challenges. The limitations noted in that study are like ours including small sample sizes, limited language scope, and evaluations conducted by healthcare professionals rather than patients. These similarities emphasise the need for further research involving diverse populations and languages to fully assess the effectiveness of GenAI-generated patient information in healthcare settings.

### 4.4 Implications for Practice

The use of GenAI-generated videos presents a promising avenue for enhancing patient communication in imaging and treatment involving radiation. Healthcare providers can leverage these tools to deliver personalised, multilingual information, improving accessibility for patients with language barriers or low literacy levels. Implementing GenAI videos in clinical practice could lead to increased patient engagement, better understanding of medical procedures, and higher satisfaction with care.

To effectively integrate these tools, practitioners should consider collaborating with language experts to refine translations and ensure cultural appropriateness. Additionally, investing in high-quality recordings and exploring GenAI platforms with advanced customisation options can enhance the naturalness of the avatar’s delivery. Addressing technical challenges, such as improving green screen keying and avatar movement, will further optimise the effectiveness of these videos.

### 4.5 Limitations of the Study

This study has several limitations that need to be considered. First, the small sample size and focus on a single language (Thai) limit the generalizability of the findings to other populations and linguistic contexts. The results may not apply to speakers of other languages or diverse cultural settings. Second, the assessment was conducted with health professionals (medical physicists and students) rather than patients, who are the ultimate end-users of this technology. Therefore, the findings may not fully reflect patient perspectives or needs. Third, scripted content might not capture the nuances and spontaneity of natural patient interactions, which are crucial for effective communication in healthcare. Finally, the study utilised only one GenAI platform (HeyGen), which may not fully represent the capabilities or limitations of other GenAI tools or future versions of the same platform. Technical factors, such as the quality of the initial avatar recording, could have influenced the results, suggesting that these aspects play a significant role in the effectiveness of GenAI-generated content.

These limitations highlight the need for caution when interpreting the results, as they may not be universally applicable across different GenAI tools, patient populations, languages, or healthcare settings.

### 4.6 Recommendations for Future Research

Future studies should test GenAI-generated videos with diverse patient populations, including individuals with varying literacy levels, age groups, and cultural backgrounds. Additionally, expanding the range of languages and dialects will be crucial to assess the scalability and adaptability of GenAI tools for global healthcare use.

Longitudinal studies could provide insights into the long-term impact of these tools on patient outcomes, such as reducing anxiety, improving treatment adherence, and enhancing overall satisfaction with care. Future research should directly explore patient perceptions and experiences, offering a more comprehensive understanding of GenAI’s practical utility and acceptability in real-world healthcare settings.

Investigating multiple GenAI platforms can help evaluate technological advancements. Additionally, addressing ethical considerations, including patient consent, data privacy, and the responsible use of AI, will be essential as these tools will likely become more integrated into clinical practice.

## 5 Conclusion

Our pilot study demonstrates the potential of GenAI as an effective tool for delivering personalised, multilingual patient communication in imaging and treatment involving radiation, specifically using the Thai language. By leveraging GenAI to create personalised videos, the research highlights the technology’s capacity to bridge communication gaps for patients facing literacy or language barriers. Despite being conducted in a single language, the findings provide valuable insights that can inform future research and development, suggesting that with further refinement and expansion to additional languages and settings, GenAI could be integrated into clinical practice to enhance patient education, engagement, and overall healthcare access and outcomes.

## Data Availability

All data produced in the present study are available upon reasonable request to the authors

## Statements & Declarations

### Ethics approval

This is an observational study. The author’s Institute Research Ethics Committee has confirmed that no ethical approval is required.

### Consent to participate

Informed consent was obtained from all individual participants surveyed in the study.

## Appendix A HeyGen Still Avatar instructions

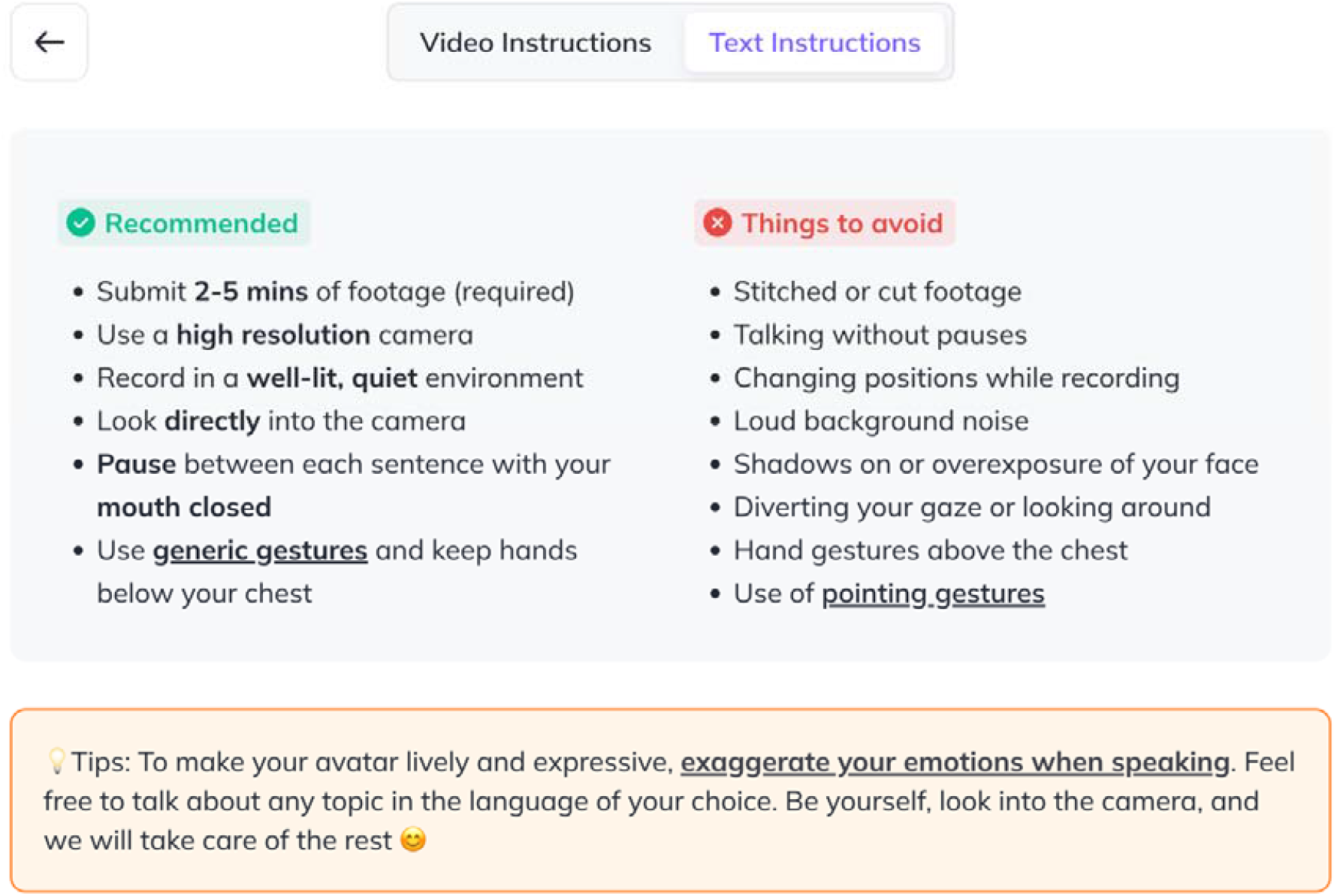

## References

1. Ishikawa H, Yano E, Fujimori S, Kinoshita M, Yamanouchi T, Yoshikawa M, et al. Patient health literacy and patient-physician information exchange during a visit. Family Practice. 2009;26:517–23.

2. Suhonen R, Nenonen H, Laukka A, Välimäki M. Patients’ informational needs and information received do not correspond in hospital. Journal of Clinical Nursing. 2005;14:1167–76.

3. Alawad S, Abujamea A. Awareness of radiation hazards in patients attending radiology departments. Radiat Environ Biophys. 2021;60:453–8.

4. Chatterjee A, Strong G, Meinert E, Milne-Ives M, Halkes M, Wyatt-Haines E. The use of video for patient information and education: A scoping review of the variability and effectiveness of interventions. Patient Education and Counseling. 2021;104:2189–99.

5. Ganguli I, Sikora C, Nestor B, Sisodia RC, Licurse A, Ferris TG, et al. A Scalable Program for Customized Patient Education Videos. The Joint Commission Journal on Quality and Patient Safety. 2017;43:606–10.

6. Schooley B, San Nicolas-Rocca T, Burkhard R. Patient-Provider Communications in Outpatient Clinic Settings: A Clinic-Based Evaluation of Mobile Device and Multimedia Mediated Communications for Patient Education. JMIR mHealth uHealth. 2015;3:e2.

7. Jlala HA, French JL, Foxall GL, Hardman JG, Bedforth NM. Effect of preoperative multimedia information on perioperative anxiety in patients undergoing procedures under regional anaesthesia. British Journal of Anaesthesia. 2010;104:369–74.

8. Bånnsgård M, Nouri A, Finizia C, Engström M, Moreno J, Roos U, et al. Communicating Patient Safety Information Through Video and Oral Formats—A Comparison. J Patient Saf. 2023;19:137–42.

9. Lasater L, Mehler PS. The Illiterate Patient: Screening and Management. Hospital Practice. 1998;33:163–70.

10. Weiss BD, Hart G, Pust RE. The Relationship Between Literacy and Health. hpu. 1991;1:351–63.

11. Habib T, Nair A, Von Pressentin K, Kaswa R, Saeed H. Do not lose your patient in translation: Using interpreters effectively in primary care. S Afr Fam Pract [Internet]. 2023 [cited 2024 Oct 17];65. Available from: https://safpj.co.za/index.php/safpj/article/view/5655

12. Patil S, Davies P. Use of Google Translate in medical communication: evaluation of accuracy. BMJ. 2014;349:g7392–g7392.

13. Taylor B, McLean G. Exploring the use of mobile translation applications for culturally and linguistically diverse patients during medical imaging examinations in Australia – a systematic review. J of Medical Radiation Sci. 2024;71:432–44.

14. Leiker D, Gyllen AR, Eldesouky I, Cukurova M. Generative AI for learning: Investigating the potential of synthetic learning videos [Internet]. arXiv; 2023 [cited 2024 Oct 17]. Available from: https://arxiv.org/abs/2304.03784

15. Chu SN, Goodell AJ. Synthetic Patients: Simulating Difficult Conversations with Multimodal Generative AI for Medical Education [Internet]. arXiv; 2024 [cited 2024 Oct 17]. Available from: https://arxiv.org/abs/2405.19941

16. Eriksen NS, Al-Bakri M, Boysen KB, Klefter ON, Schmidt DC, Reinwaldt K, et al. Generative artificial intelligence for increasing accessibility of patient information videos in ophthalmology. AJO International. 2024;1:100016.

17. Papineni K, Roukos S, Ward T, Zhu W-J. BLEU: a method for automatic evaluation of machine translation. Proceedings of the 40th Annual Meeting on Association for Computational Linguistics - ACL ‘02 [Internet]. Philadelphia, Pennsylvania: Association for Computational Linguistics; 2001 [cited 2024 Nov 4]. p. 311. Available from: http://portal.acm.org/citation.cfm?doid=1073083.1073135

